# Curation of Mini Mental State Examination (MMSE) Scores in the VA Million Veteran Program (MVP): Applications for Cognitive Aging Research

**DOI:** 10.64898/2026.07.14.26358064

**Authors:** Francesca V. Lopez, Margaret Gillis, Sophia Lee, McKenna S. Sakamoto, Rui Zhang, VA Million Veteran Program, Richard Sherva, Mark W. Logue, Victoria C. Merritt

## Abstract

**Background:** Electronic health record (EHR)-linked biorepositories provide opportunities to advance epidemiological research in Alzheimer’s disease (AD) and related dementias.

**Objective:** Evaluate the extraction, curation, and associative validity of Mini Mental State Examination (MMSE) scores from the VA EHR for participants in the VA Million Veteran Program (MVP).

**Methods:** The sample (N = 49,555; 7.4% women) included a multiethnic cohort (European [68.3%], African [20.4%], Hispanic [9.0%]) with EHR-extracted MMSE scores; 30.7% were apolipoprotein E (*APOE*) ε4 carriers, and 25.8% had multiple scores. Linear regressions examined cross-sectional associations between ε4 dosage (0, 1, 2) and first and lowest MMSE scores. MMSE scores were also evaluated against MVP dementia diagnostic algorithms in participants aged ≥65 years.

**Results:** Among participants of European ancestry, there was a significant ε4 dose-response relationship (*p*s < .001) with MMSE scores. Homozygote carriers scored lower than heterozygote carriers (M_diff_: first = -0.5; lowest = -0.9), who scored lower than non-carriers (M_diff_: first = -0.4; lowest = -0.6). Among Veterans of African and Hispanic ancestry, no dose-response relationship was observed, although ε4 carriers had lower scores than non-carriers (*p*s ≤ .04). MMSE scores corresponded strongly with dementia case/control status across phenotypes: mild impairment on the MMSE was strongly associated with AD (odds ratio [OR] = 11.48), with more severe MMSE impairment showing stronger associations (moderate OR = 17.95; severe OR = 27.83).

**Conclusion:** This study demonstrated MMSE scores can be systematically extracted and curated from the VA EHR. Findings offer a scalable framework for future studies on risk stratification, highlighting the potential for harnessing MVP to explore genetic and clinical factors contributing to cognitive and dementia outcomes in diverse samples.

## Introduction

Alzheimer’s disease (AD) and related dementias (ADRD) present a significant and growing challenge to public health. As of 2024, more than 7 million older adults in the United States (U.S.) and 40 million globally were estimated to live with AD/ADRD. These numbers are projected to increase substantially over the next several decades^1^. The economic burden is equally significant with annual healthcare costs exceeding $350 billion in the U.S. and $1.5 trillion worldwide^2^. Despite this, our understanding of the genetic, environmental, and clinical mechanisms contributing to ADRD remains incomplete. Among genetic risk factors, the apolipoprotein (*APOE*) ε4 allele remains the strongest and most well-established genetic risk factor for late-onset AD, with each ε4 allele associated with increased AD risk in a dose-dependent manner ^3–5^. Large-scale, real-world datasets are critical for advancing our understanding of ADRD risk and developing effective intervention and prevention strategies^3^.

Electronic health records (EHRs) provide a rich source of real-world clinical information, but extracting reliable cognitive data at scale remains a significant challenge. Objective cognitive assessments are critical for identifying persons at greatest risk for future cognitive decline and ADRD. Due to demands on neuropsychological services, brief cognitive screeners are typically used as a viable first-line assessment option. Ideal screeners are efficient to administer, assess multiple cognitive domains, and identify individuals at risk for decline^6^. The Mini Mental State Examination (MMSE^7^) exemplifies one of the most widely used screeners in clinical and research settings with its extensive historical use in VA clinical practice that positions it as the most consistently available cognitive measure within the VA EHR. While the MMSE was designed as a screening tool rather than a diagnostic instrument, its widespread clinical adoption and validation across different populations establish it as a valuable marker of global cognition for large-scale epidemiologic research^8^.

However, the heterogeneity and unstructured nature of clinical data, where key data are often incomplete or inconsistently documented, have been a major barrier. While natural language processing and machine learning show promise^9–13^ for extracting cognitive data at scale, the variability in how scores are documented in free-text clinical notes (e.g., inconsistent formatting, multiple candidate values within a single note, non-standard or modified test administrations) raises the question of whether automated extraction alone can reliably distinguish valid from invalid entries. As such, automated methods may require rigorous validation and expert review to ensure accuracy and reliability^14^. High-quality, objective cognitive data would allow for the direct examination of cognitive status over time as well as relationships with key risk factors (e.g., genetics). Since genetic analyses were designed to test dose-response relationships between *APOE* and cognitive performance, a continuous outcome measure was required. Accordingly, we developed an extraction approach that combines automated-rule-based extraction with manual curation to evaluate whether these data capture expected genetic and clinical patterns relevant in cognitive aging and AD/ADRD.

The Veterans Affairs (VA) Million Veteran Program (MVP) is uniquely positioned to support large-scale extraction and curation of cognitive data from the EHR, and to enhance our understanding of cognitive decline and ADRD risk in real-world populations. As one of the largest EHR-biorepositories to date, MVP includes genomic, clinical, and survey data from over one million U.S. Veterans^15^. The MVP Cognitive Working Group has leveraged these resources (i.e., diagnostic codes from the EHR, genomic profiles, survey data, or their combinations) to advance our understanding of ADRD risk^16–21^. The existing infrastructure (i.e., genome-wide genetic data, validated dementia diagnostic algorithms across a large multi-ethnic cohort, linked EHR access) makes MVP an ideal platform for extracting and evaluating objective cognitive performance data (i.e., MMSE scores) and examining their relationships with established genetic and clinical markers of AD/ADRD risk. Building on this work, the current study aimed to develop and evaluate a scalable method for extracting cognitive data from the EHR using MVP’s infrastructure. We first characterized our cohort (i.e., Veterans with EHR-curated MMSE scores) by examining pertinent demographics, providing key reference information for future MVP studies of cognitive aging and ADRD. Additionally, we evaluated relationships between EHR-curated MMSE scores and *APOE* ε4 allele. Finally, we examined relationships between MMSE scores and four MVP-validated dementia phenotypes^21^. This multi-layered approach (i.e., characterizing the cohort, evaluating associations with genetic risk factors, evaluating associations with dementia phenotypes) was designed to assess the associative validity of curated cognitive data from the VA EHR across genetic and clinical domains. First, we hypothesized that the associative validity of EHR-curated MMSE scores would be evidenced by a dose-dependent inverse relationship with *APOE* ε4 dosage and AD polygenic risk scores. Second, we hypothesized that the magnitude of these relationships would vary as a function of ancestral background given known differences in *APOE* ε4 penetrance across European, African, and Hispanic ancestry groups.

## Methods

### Million Veteran Program (MVP)

The current study used data from MVP, a VA research initiative developed over a decade ago that examines genetic and environmental contributions to Veteran health and well-being. At present, MVP is one of the largest national databases of health and genetic information in the world^15^. MVP received VA Central Institutional Review Board (cIRB) approval in 2010, and the present study – conducted under MVP projects “MVP015” and “MVP040” – received institutional oversight approval in 2018 and 2022, respectively.

Initially, any Veteran enrolled in the Veterans Health Administration (VHA) was eligible to enroll in MVP, but eligibility was recently opened to the entire Veteran population, regardless of VHA-user status. MVP-enrolled Veterans provided written informed consent and agreed to allow MVP investigators to access data from their EHR. Upon enrollment, Veterans were asked to complete MVP self-report measures assessing a wide range of sociodemographic, military, and health factors^22^, as well as provide a blood sample for genetic analysis. Detailed information about the study design and cohort characteristics are reported in Gaziano et al.^15^. This study used MVP data release v20_1. *Cognitive Data Extraction and Curation*

We selected MMSE total scores rather than the individual subdomain scores as the outcome of interest for two primary reasons. First, the MMSE is the most commonly administered cognitive screener in the VA based on our examination of the VA EHR, and much more common than full neuropsychological testing. Second, the validity and reliability of the MMSE total score as a screener for global cognitive function is well-established, whereas the individual subdomains have not demonstrated sufficient validity or reliability to capture their intended domains^23^. Furthermore, within the VA EHR, MMSE scores are not consistently captured in structured, discrete data fields; instead, these scores are documented within free-text clinical notes (e.g., progress notes, neuropsychology evaluations, dementia screening notes). As such, extraction of MMSE scores required text-processing approaches applied to unstructured note data as described below. Given these documentation patterns, prioritizing total scores maximized feasibility and data quality for large-scale extraction.

The MVP Data Core extracted cognitive data (i.e., MMSE scores) from unstructured data sources (i.e., free-text clinical notes) within the EHR. Specifically, structured queries and rule-based algorithms were used to isolate MMSE score entries from the clinical note narrative text. The following key terms were used as search terms in this process: “MMSE,” “Mini Mental State Examination,” “Dementia Screening,” and “Cognitive Impairment Screening”. The selected terms reflect common clinical documentation conventions for MMSE reporting within VA clinical notes and were used to maximize sensitivity for identifying relevant data. Ninety characters surrounding each key term (referred to subsequently as the “text snippet”) were extracted from the EHR, along with the associated note date and patient ID. This window size was selected to balance the inclusion of relevant numeric values with the exclusion of unrelated numerical content. This yielded 1,165,112 records from 92,921 unique participants. Duplicate records were removed (N=873,835 records), resulting in 291,277 available records from 92,921 unique participants.

After patient-identifier scrubbing by the MVP Data Core, two candidate scores were extracted from each text snippet using complementary rule-based strategies: (1) the first positive number after the key term (“Score 1”) and (2) the first number appearing within the 90-character text snippet, allowing for capture of values that may precede the key term (“Score 2”). Often, when there were no numbers preceding the key term, these were the same score. These two extraction strategies were used because MMSE scores were documented inconsistently relative to the key term; for example, in some notes, the score appeared immediately after the term “MMSE” (e.g., “MMSE: 24/30”), while in other notes, the score was preceded by other numeric values (e.g., dates, vital signs, prior scores) within the same text snippet. Extracting both candidate values allowed the manual review process to identify the correct score when the two diverged. These data (i.e., Score 1 and Score 2) were then compiled into a Microsoft Access template for review. The template also included the note date, patient ID, and the 90-character text snippet. Additionally, there were columns in the template with checkboxes for the reviewers to indicate the following: whether Score 1 was correct; whether Score 2 was correct; or whether no valid score existed in the 90-character text snippet. A final column included a cell for manual entry of the MMSE score if there was a valid score within the text snippet that was not reflected in Score 1 or Score 2.

Entries where Score 1 was an integer from 0-30 followed by “/30” were flagged as “likely correct” (records = 104,563) based on proper range and common MMSE notation, excluding any date-like patterns (e.g., “12/30/2010”). Next, all scores not flagged as “likely correct” (records = 186,714/291,277; 64% of the available records) underwent individual manual review by trained reviewers following a standardized protocol. Following standardized training, a total of five reviewers were assigned non-overlapping batches of records and were instructed to review the text snippet for each entry (i.e., row) in the Microsoft Access template to determine whether there was a valid MMSE score present and then to mark the appropriate cells (as indicated above). Reviewers were directed to mark “not valid” for any of the following situations: when an alternate or modified version of the MMSE was used (e.g., MMSE total score was not out of 30 [“/30”]); implausible scores (e.g., MMSE total score > 30); or scores with other mitigating factors were indicated in the text (e.g., intoxication). When multiple scores appeared in the same 90-character text snippet, reviewers selected the MMSE score from the test administered closest in time to the note date.

Since each record was reviewed by a single reviewer, formal disagreement resolution procedures were not carried out, and inter-rater reliability metrics (e.g., Cohen’s kappa) were not calculated as part of the primary curation workflow. However, we conducted a supplemental reliability analysis on a random subset of 1000 records reviewed by three independent reviewers (SB, VCM, MWL). Overall agreement was excellent (κ = .968 for identification of a valid versus invalid score; κ = .994 for records with a valid numeric score; κ = .958 across the full sample including non-valid entries).

After review, “likely correct” and manually screened scores were assembled and range checked. Exceptionally low scores (e.g., < 10) were manually verified through note text examination. Given that clinical notes often contain copied text and references to prior test scores, another round of data deduplication occurred, retaining only the first instance of each unique score per participant to reduce redundancy from repeated documentation of prior test results in clinical notes. After this final round of data deduplication, 83,664 records from 59,001 unique participants remained. Of these, 49,555 had available genetic data and were included in all analyses as *APOE* ε4 was the primary predictor of interest. Missingness was minimal and related to *APOE* ε4 genotype data. Among participants with curated MMSE scores, 236 were missing *APOE* ε4 genotype data. For analyses examining ε4 dosage, 77 participants of European ancestry, 133 participants of African ancestry, and 10 participants of Hispanic ancestry were excluded; an additional 16 participants with missing *APOE* ε4 genotype data were excluded in the ancestry-specific genetic analyses. A flow chart of the full extraction and curation process is shown in Supplemental Figure 1.

### Genetics Data

Genotype data processing and cleaning was performed by the MVP Bioinformatics Core. The chip design and genotyping cleaning pipeline have been described elsewhere^24^. *APOE* ε4 genotypes were generated based on the imputed genotypes for the two single nucleotide polymorphisms (SNPs) used to determine the isoform: rs7412 and rs429358. SNP data were extracted from the Phase 4 MVP genotype release, which includes data for approximately 650,000 MVP participants. Imputation was performed using the NHLBI Trans-Omics for Precision Medicine (TOPMed) reference panel. Both rs7412 and rs429358 were well imputed across ancestry groups, with imputation quality of r^2^ > 0.99 in participants of European and Hispanic ancestry, and r^2^ > 0.98 in participants of African ancestry. The “best-guess” genotypes for these SNPs were generated from the imputed data using a certainty threshold of 90%. To ensure that any genetic associations were not limited to *APOE* ε4 dosage, an AD polygenic risk score (PRS) was calculated^25^. This PRS excluded the *APOE* region on chromosome 19 and is, therefore, an index of AD genetic risk summarizing the contribution from other known risk loci throughout the genome. Genetic analyses relied on the harmonized ancestry and race/ethnicity (HARE) method^26^ which uses a combination of self-reported ancestry and genotype data. The

HARE method classifies individuals into four groups: European ancestry, African ancestry, Hispanic ancestry, and Asian ancestry. Ancestry-specific principal components (PCs) were computed in several steps. First, we separated participants based on their HARE assignments. Then we performed linkage disequilibrium (LD) pruning on the genotyped SNP data with a window size of 50 variants, a step size of 5 variants, and an r^2^ threshold of 0.2, Finally, we performed principal component analysis (PCA) using FlashPCA.

### MVP Dementia-Related Diagnostic Algorithms

As described elsewhere^21^, we developed four phenotypes related to AD and dementia using ICD codes adapted to the difficulties of identifying AD cases based on the limitations of the VA EHR. From most narrow to most broad, our ICD code-based phenotypes were: (1) “Strict AD”, which includes AD-specific ICD codes only; (2) “AD+,” which includes AD and other non-specific dementia codes (e.g., dementia without behavioral disturbance); (3) Alzheimer’s disease and related dementias (“ADRD”), which includes AD and related dementias such as vascular dementia; and (4) all-cause “dementia.” These four diagnosis levels are nested by design. In other words, if an MVP participant is classified as an AD case, then the same participant also meets criteria for AD+, ADRD, and dementia. Cases were defined as having a disease onset (first dementia ICD code) age of 65 or older. Additionally, at least two qualifying ICD codes (on different dates) were required to meet the “case” definition. Controls were defined as participants who did not meet criteria for any of the four dementia phenotypes (i.e., no ICD codes for dementia), had no ICD codes for mild cognitive impairment, and had no record of AD-related medications. Controls were used as a common comparison group across all phenotype analyses. The four diagnostic phenotypes described above represent operational definitions of AD/ADRD at varying levels of diagnostic specificity ranging from the most restrictive (“Strict AD”) to the most inclusive (all-cause “dementia”). Throughout this manuscript, AD/ADRD is used to broadly refer to the disease-specific process consistent with the use in the Introduction and Discussion, while “Strict AD,” “AD+,” “ADRD,” and “dementia” (in quotation marks) refer to these specific operational phenotypes.

### Analyses

Descriptive statistics were used to summarize the overall study cohort in terms of demographic characteristics (i.e., age, sex, genetic ancestry) as well as availability and distribution of MMSE scores. Two MMSE total scores were used: (1) first MMSE score recorded to approximate baseline cognitive status, and (2) lowest MMSE score recorded to capture the greatest documented cognitive impairment over time. Age was determined at time of MMSE assessment.

Linear regression models were used to examine relationships between MMSE scores and genetic risk factors (i.e., *APOE* ε4, AD PRS). For *APOE* ε4 dosage, the number of ε4 alleles (0, 1, or 2) was coded for analyses. Tukey corrected pairwise comparisons were used to examine simple effects. Models were stratified by genetic ancestry and adjusted for age, sex, and the first 3 genetic ancestry PCs. Effect sizes (*R*_P_^2^), 95% confidence intervals (CI), and established levels of statistical significance were used to guide interpretation.

The final set of analyses were restricted to participants aged ≥ 65 years (at first MMSE assessment) given this group’s increased risk for ADRD and greater relevance to clinical decision-making involving the MMSE. Established MMSE cutoff scores were applied to categorize severity of cognitive impairment: mild impairment ≤ 25, moderate impairment ≤ 20, and severe impairment ≤ 10^27^. For each cutoff and phenotype, a contingency table evaluated the performance of the lowest MMSE score as an indicator of dementia presence/absence against each MVP dementia-related diagnostic algorithm. Sensitivity (true positive rate; frequency of dementia cases that met the MMSE impairment threshold), specificity (true negative rate; proportion of controls that did not meet the threshold), with 95% CI, evaluated diagnostic accuracy of MMSE thresholds across dementia phenotypes. Specificity is identical for all dementia phenotypes as all used similarly defined control sets.

### MVP Cohort Sample Characteristics

The study sample included 49,555 MVP participants with available genetic, EHR, and curated MMSE total scores. The cohort was multiethnic: European (68.3%) African (20.4%), Hispanic (9.0%), and Asian (0.8%). Population coverage of curated MMSE scores varied across ancestry groups with MMSE data available for 7.28% of participants of European ancestry, 8.23% of African ancestry, 8.53% of participants of Hispanic ancestry, and 4.93% of participants of Asian ancestry. The number of participants with available MMSE data in the Asian ancestry group was insufficient to support reliable ancestry-stratified analyses; thus, this ancestry group was excluded from all analyses. Approximately 30.7% were *APOE* ε4 carriers, and 25.8% had more than one MMSE score. See Table 1 for cohort characteristics and availability and distribution of MMSE scores.

**Table 1.**
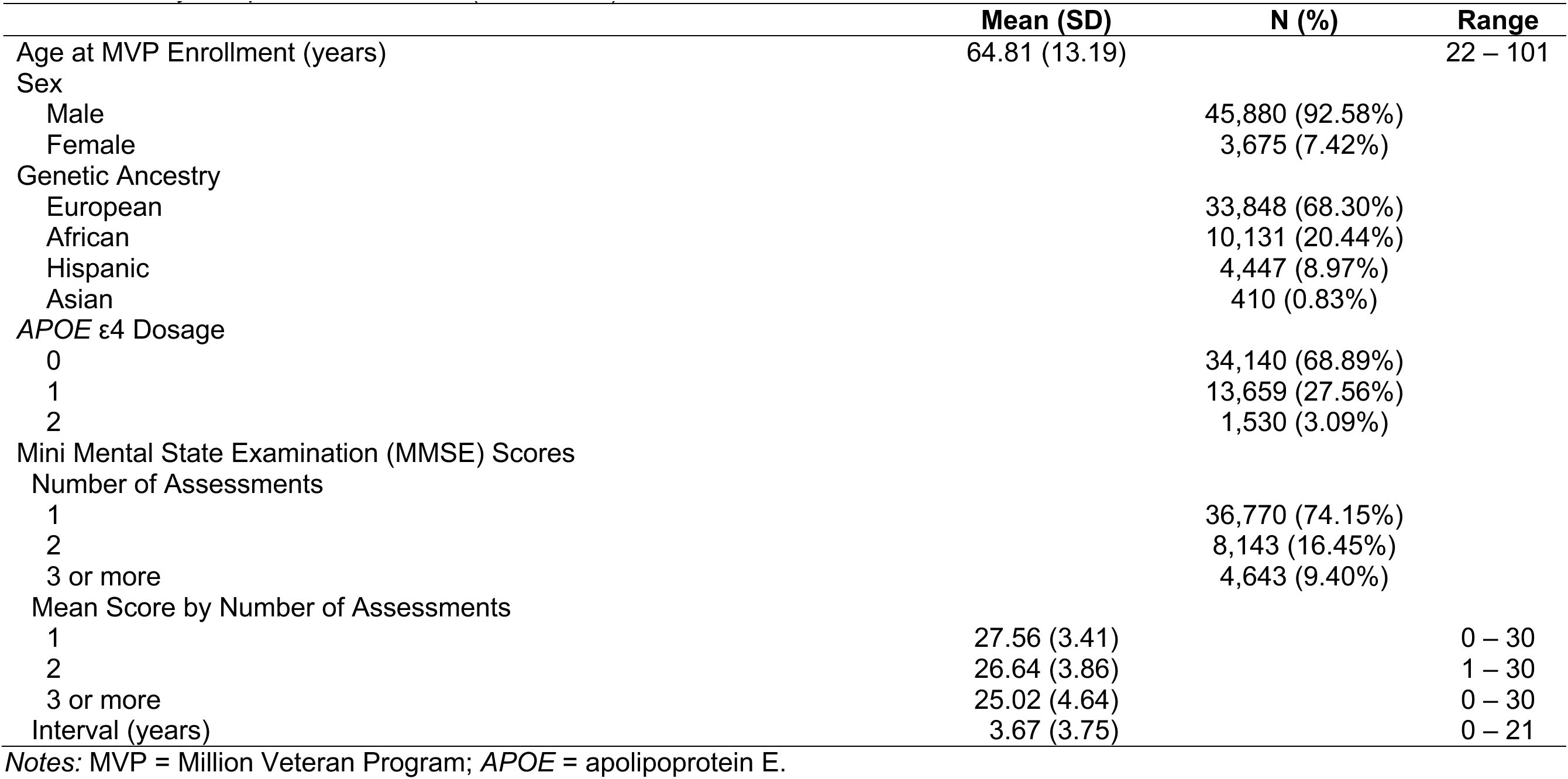
Study sample characteristics (N = 49,555).

## Results

### MMSE Associations with APOE ε4 Genotype & AD PRS

Among participants of European ancestry, there was a significant dose-response relationship between *APOE* ε4 genotype and MMSE total scores (*p*s < .001; *R*_P_^2^ = 0.005-0.008; Table 2). On average, homozygote ε4 carriers performed lower than heterozygote ε4 carriers, who performed lower than non-carriers for both first and lowest MMSE total scores (*p*s < .001; Figure 1). In European ancestry participants, higher AD PRS scores were also associated with lower MMSE total scores (*p*s < .001).

**Table 2.** Multiple Linear Regressions between *APOE* ε4 Genotype with MMSE Total Scores.

| Predictor | First MMSE Score<br>(B, 95% CI) | SE | *p* | Lowest MMSE Score<br>(B, 95% CI) | SE | *p* |
| --- | --- | --- | --- | --- | --- | --- |
| *European Ancestry* |  |  |  |  |  |  |
| *APOE* ε4 Genotype | -0.14 (-0.15, -0.12) | 0.01 | <.001 | -0.17 (-0.19, -0.15) | 0.01 | <.001 |
| Sex | -0.07 (-0.11, -0.03) | 0.02 | <.001 | -0.05 (-0.09, 0.00) | 0.02 | .030 |
| Age | -0.26 (-0.27, -0.25) | 0.01 | <.001 | -0.30 (-0.31, -0.29) | 0.01 | <.001 |
| European Ancestry PC1 | 0.02 (0.01, 0.03) | 0.01 | <.001 | 0.02 (0.01, 0.03) | 0.01 | <.001 |
| European Ancestry PC2 | 0.00 (-0.01, 0.01) | 0.01 | .450 | 0.00 (-0.01, 0.01) | 0.01 | .530 |
| European Ancestry PC3 | -0.02 (-0.03, -0.01) | 0.01 | .010 | -0.02 (-0.03, -0.01) | 0.01 | <.001 |
| *African Ancestry* |  |  |  |  |  |  |
| *APOE* ε4 Genotype | -0.07 (-0.09, -0.03) | 0.02 | <.001 | -0.08 (-0.11, -0.05) | 0.03 | <.001 |
| Sex | -0.15 (-0.22, -0.09) | 0.03 | <.001 | -0.14 (-0.20, -0.08) | 0.01 | <.001 |
| Age | -0.30 (-0.32, -0.28) | 0.01 | <.001 | -0.35 (-0.38, -0.34) | 0.03 | <.001 |
| African Ancestry PC1 | 0.02 (-0.01, 0.05) | 0.01 | <.001 | 0.02 (-0.00, 0.03) | 0.01 | .100 |
| African Ancestry PC2 | -0.00 (-0.02, 0.02) | 0.01 | .790 | 0.01 (-0.01, 0.03) | 0.01 | .260 |
| African Ancestry PC3 | -0.01(-0.02, 0.01) | 0.01 | .400 | 0.00 (-0.02, 0.02) | 0.01 | .780 |
| *Hispanic Ancestry* |  |  |  |  |  |  |
| *APOE* ε4 Genotype | -0.10 (-0.16, -0.04) | 0.03 | <.001 | -0.13 (-0.19, -0.07) | 0.03 | <.001 |
| Sex | -0.05 (-0.16, 0.06) | 0.05 | .400 | -0.06 (-0.18, 0.05) | 0.06 | .280 |
| Age | -0.31 (-0.33, -0.28) | 0.01 | <.001 | -0.36 (-0.39, -0.33) | 0.01 | <.001 |
| Hispanic Ancestry PC1 | 0.05 (0.02, 0.07) | 0.01 | <.001 | 0.03 (0.00, 0.06) | 0.01 | .030 |
| Hispanic Ancestry PC2 | 0.02 (-0.01, 0.05) | 0.01 | .150 | 0.02 (-0.01, 0.05) | 0.01 | .170 |
| Hispanic Ancestry PC3 | 0.01 (-0.01, 0.04) | 0.01 | .350 | 0.02 (-0.01, 0.04) | 0.01 | .250 |

Notes: *APOE* = apolipoprotein E; MMSE = Mini Mental State Examination; CI = confidence interval; SE = standard error; PC = principal components.

In African and Hispanic ancestry participants, we also observed associations between *APOE* ε4 genotype and MMSE scores, with *p*s < .001; *R*_P_^2^ = 0.002-0.003 in African ancestry participants and *p*s < .001; *R*_P_^2^ = 0.002-0.004 in Hispanic ancestry participants. On average, *APOE* ε4 carriers performed lower than non-carriers for both first and lowest MMSE total scores (*ps* ≤ .04), but no significant dose-response relationship was observed between homozygote and heterozygote ε4 carriers (Figure 1) in either the African or Hispanic ancestry analyses.

### MMSE Associations with Dementia-Diagnostic Algorithms

MMSE scores were strongly associated with dementia case status across all MMSE severity thresholds and diagnostic phenotypes, with lower MMSE thresholds (i.e., greater cognitive impairment) corresponding to substantially higher odds of having a dementia diagnosis. See Table 3. For example, as the MMSE threshold was lowered, the odds of dementia increased: participants meeting the mild (MMSE ≤ 25), moderate (MMSE ≤ 20), and severe (MMSE ≤ 10) thresholds had 11-, 18-, and 28-fold higher odds of being Strict AD cases. The odds ratios followed a similar pattern across the different dementia diagnoses, but with lower observed odds ratios as the dementia diagnosis was expanded from Strict AD to dementia. The specificity of these cognitive impairment thresholds was high: 85%, 97%, and 100% of the lowest observed MMSE scores for controls were above the mild, moderate, and severe thresholds, respectively.

**Table 3.** Sensitivity and Specificity of MMSE Impairment Thresholds by Dementia-Related Phenotypes.

| Impairment Threshold | AD |  |  | AD+ |  |  | ADRD |  |  | Dementia |  |  |
| --- | --- | --- | --- | --- | --- | --- | --- | --- | --- | --- | --- | --- |
|  | OR | Sensitivity | Specificity | OR | Sensitivity | Specificity | OR | Sensitivity | Specificity | OR | Sensitivity | Specificity |
| Mild (MMSE≤25) | 11.48<br>(10.30,12.81) | 0.67<br>(0.65,0.69) | 0.85<br>(0.84,0.86) | 8.17<br>(7.49, 8.93) | 0.59<br>(0.58,0.60) | 0.85<br>(0.84,0.86) | 7.70<br>(7.07,8.40) | 0.58<br>(0.57,0.59) | 0.85<br>(0.84,0.86) | 6.68<br>(6.14,7.28) | 0.54<br>(0.53,0.55) | 0.85<br>(0.84,0.86) |
| Moderate (MMSE≤20) | 17.95<br>(15.05,21.53) | 0.35<br>(0.34,0.37) | 0.97<br>(0.97,0.97) | 12.43<br>(10.52,14.75) | 0.28<br>(0.27,0.29) | 0.97<br>(0.97,0.97) | 11.64<br>(9.86,13.81) | 0.26<br>(0.25,0.27) | 0.97<br>(0.97,0.97) | 10.33<br>(8.76,12.25) | 0.24<br>(0.23,0.25) | 0.97<br>(0.97,0.97) |
| Severe (MMSE≤10) | 27.83<br>(15.11,57.04) | 0.05<br>(0.05,0.06) | 1.00<br>(1.00,1.00) | 17.27<br>(9.50,35.04) | 0.03<br>(0.03,0.04) | 1.00<br>(1.00,1.00) | 16.25<br>(8.94,32.91) | 0.03<br>(0.03,0.03) | 1.00<br>(1.00,1.00) | 14.56<br>(8.01,29.49) | 0.03<br>(0.03,0.03) | 1.00<br>(1.00,1.00) |

*Notes:* MMSE = Mini Mental State Examination; OR = Odds Ratio. From most narrow to most broad, our ICD code-based phenotypes were: (1) “Strict AD”, which includes AD-specific ICD codes only; (2) “AD+,” which includes AD and other non-specific dementia codes (e.g., dementia without behavioral disturbance); (3) Alzheimer’s disease and related dementias (“ADRD”), which includes AD and related dementias such as vascular dementia; and (4) all-cause “dementia.”

## Discussion

This study makes two related contributions: (1) a scalable, structured approach for curating cognitive data from the VA EHR, and (2) evidence for the associative validity of the resulting scores against established genetic and clinical correlates of AD/ADRD. We evaluated curated scores against two domains in a sample of nearly 50,000 Veterans: (1) AD genetic risk and (2) MVP-curated dementia diagnostic algorithms. We reproduced well-established genetic and clinical patterns, providing support for our approach for extracting and incorporating objective cognitive data in MVP.

We observed clear ancestry-specific *APOE* ε4 effects. Among participants of European ancestry, there was a robust dose-response relationship with higher *APOE* ε4 dosage related to progressively lower MMSE scores. In comparison, both African and Hispanic ancestry participants exhibited reduced allelic penetrance. Although ε4 carriers had lower MMSE scores than non-carriers, the dosage gradient was attenuated, and the heterozygote-homozygote differences were not statistically distinguishable in our sample. These patterns align with prior epidemiological and genetic studies documenting relationships between *APOE* ε4 and cognitive outcomes across ancestry groups^3, 28–29^. Some studies report variable ε4 penetrance^30–31^, while others report intermediate heterozygote effects^32–36^ in African and Hispanic ancestry samples. These inconsistencies likely reflect differences in sample size, admixture proportion, and age across cohorts. Together with the significant AD PRS association, these findings suggest that EHR-curated MMSE scores capture meaningful cognitive variation and are linked with well-established AD genetic risk factors. While the proportion of variance explained was modest and the observed mean differences in MMSE scores were small in absolute terms, effect sizes of this magnitude are consistent with those reported in large population-based studies of genetic associations with cognitive outcomes, and small mean differences at the population level can carry meaningful implications for identifying persons at elevated dementia risk^37–38^. Importantly, the replication of these nuanced, ancestry-specific patterns highlights both the complexity of genetic effects across populations, and the value of a large, multiethnic biorepository such as MVP for biomarker validation and risk stratification studies.

Beyond genetic relationships, MMSE scores demonstrated strong concordance with MVP-curated dementia diagnostic algorithms, with odds ratios of 7- to 28-fold across all thresholds and phenotypes. Participants with *greater* cognitive impairment had substantially higher odds of being a dementia case. Another notable finding was that regardless of the impairment threshold, the odds of being a dementia case diminished as the dementia phenotype was broadened and made more general. For example, at the mild impairment threshold (MMSE ≤ 25), 67% of Strict AD cases showed cognitive impairment compared to 54-59% of cases using broader phenotype definitions. This pattern likely reflects that MMSE scores do not capture the full range of symptom profiles of broader (i.e., more heterogeneous) dementia classifications and are more indicative of classical AD presentation. Alternatively, this could be an indication that VA clinicians are more comfortable assigning AD-specific treatment codes to those with *very serious* levels of cognitive impairment. Those who have received MMSE scores indicative of impairment are very likely to have some indication of dementia (e.g., treatment codes for dementia, MMSE, or prescription of AD medication) based on the specificity results. Because the same control set was used for every phenotype comparison, specificity values are identical across all four dementia categories, and only the case definitions (and therefore sensitivity) vary by phenotype. Specificity was high across thresholds: of those classified as controls, only 15% had an MMSE score below the mild impairment threshold, and none had a score within the severe impairment range. These clinically relevant patterns indicate that MVP-curated MMSE scores capture meaningful cognitive variation and can be leveraged to inform cognitive phenotype definitions to optimize case identification and improve risk stratification.

These findings have several important implications. First, this study represents the first large-scale extraction and curation of cognitive data from the VA EHR within MVP. By doing so, we demonstrated that cognitive data embedded within the VA EHR can be extracted and curated at scale using a structured, protocol-driven approach. Second, we developed a scalable and methodological framework for future large-scale curation of domain-specific neuropsychological tests. The extraction pipeline is compatible with common data models and cross-system informatics architectures, which supports implementation across health systems. Third, curated MMSE scores can be integrated into multimodal research pipelines (i.e., genomics, biomarkers) to support large-scale investigations of cognitive aging and ADRD, ultimately supporting the development of risk-prediction and early detection models tailored to a variety of clinical populations. Together, findings provide a scalable framework for extracting cognitive data in large biobanks and support future epidemiological studies of ADRD. Although the current pipeline required substantial manual review, its structured and protocol-driven process is reproducible and adaptive. Future studies may benefit from increased automation as NLP methods continue to improve.

The current study is not without limitations. Our cohort was drawn from MVP participants who voluntarily enrolled and had available cognitive data. Although our sample reflected the demographic composition of the Veteran population (92.6% male, 68.3% European ancestry), findings may not generalize to the entire Veteran population (e.g., female Veterans) or the civilian population who can differ in presentation and risk factor profiles. Furthermore, the Asian ancestry group was excluded from all analyses due to insufficient sample size (n = 410), which limits the generalizability of findings to this population and underscores the need for targeted recruitment and retention efforts in future studies. Nonetheless, MVP’s scale and heterogeneity provide opportunities for future validation in external and population-based cohorts. While the MMSE is one of the most widely used cognitive screeners in clinical settings, it has known limitations including limited domain-specificity, ceiling effects, and susceptibility to educational and cultural factors, which may attenuate observed relationships. Future efforts will expand extraction to additional cognitive tests for a more granular assessment of cognitive functioning, including curation of MMSE subdomain scores, which would require more comprehensive NLP-based mining across note types. Although the manual review process served as a quality check on automated extraction with trained reviewers following a standardized protocol to adjudicate records flagged by the algorithm, records were not reviewed in duplicate across reviewers during the main curation process. We conducted a supplemental inter-rater reliability analysis on a subset of records. Overall, agreement was excellent across all comparisons, providing a strong empirical estimate of the reliability of our curation process. Still, future curation efforts may benefit from dual-reviewer protocols to further strengthen this process. Finally, our study design was cross-sectional, which limits any causal inferences that can be drawn. However, the methodological infrastructure developed here will support continued extraction and curation efforts. Future studies will examine rates of cognitive decline, predictors of progression, and the impact of risk factors across the disease course.

In summary, this study demonstrated that objective cognitive data can be systematically curated from the VA EHR at scale, leveraging the infrastructure of MVP. Our curated MMSE scores replicated known genetic and clinical relationships across three different ancestry cohorts, supporting the utility of EHR-derived cognitive data for clinical research applications. These findings establish MVP – one of the largest multiethnic biorepositories to date – as a powerful resource for integrating high-quality cognitive data into large-scale studies of cognitive aging and ADRD including future longitudinal investigations of cognitive decline and disease progression.

## Statements and Declarations

## Acknowledgments

This research is based on data from the Million Veteran Program, Office of Research and Development, Veterans Health Administration, and was supported by MVP 000 as well as awards BX004192, BX005749, IK2 CX001952, and BX006350.

This publication does not represent the views of the Department of Veteran Affairs or the United States Government. The authors sincerely thank the Veterans who volunteered to participate in the Million Veteran Program.

## Ethical Considerations

The VA Million Veteran Program (MVP) was approved by the VA Central Institutional Review Board (cIRB) in 2010 and the current studies (“MVP015;” “MVP040”) under which this project was conducted received cIRB approval in 2018 and 2022, respectively (IRB project #CIRB-18-17; #CIRB-E22-23). All Veterans provided informed consent for this study. The research was conducted in accordance with the Declaration of Helsinki.

## CRediT Authorship Contribution Statement

*Francesca V. Lopez:* Methodology, Writing – original draft.

*Margaret Gillis:* Methodology, Visualization, Formal analysis, Writing – review & editing.

*Sophia Lee:* Methodology, Writing – review and editing.

*McKenna S. Sakamoto:* Methodology, Writing – review and editing.

*Rui Zhang:* Methodology, Writing – review and editing.

*Richard Sherva*: Methodology, Software, Writing – review and editing.

*Mark W. Logue:* Conceptualization, Data curation, Funding acquisition, Investigation, Methodology, Project administration, Supervision, Writing – review and editing.

*Victoria C. Merritt:* Conceptualization, Data curation, Funding acquisition, Investigation, Methodology, Project administration, Supervision, Writing – review and editing.

## Data Availability

The data and code used to generate MVP results are accessible to researchers with MVP data access. Due to VA policy, MVP is currently only accessible to researchers with a funded MVP project (e.g., VA Merit Award, Career Development Award, NIH R01). Thus, the datasets generated and/or analyzed during the current study are not publicly available, but the corresponding author is willing to engage with reasonable requests, share code, and answer questions about the present study. Information about accessing MVP data can be found at: https://www.mvp.va.gov/pwa/joinmvp. dbGaP accession number: phs001672.

